# Space-time patterns, change, and propagation of COVID-19 risk relative to the intervention scenarios in Bangladesh

**DOI:** 10.1101/2020.07.15.20154757

**Authors:** Arif Masrur, Manzhu Yu, Wei Luo, Ashraf Dewan

## Abstract

The novel coronavirus (COVID-19) pandemic continues to be a significant public health threat worldwide. As of mid-June 2020, COVID-19 has spread worldwide with more than 7.7 million confirmed cases and more than 400,000 deaths. The impacts are substantial particularly in developing and densely populated countries like Bangladesh with inadequate health care facilities, where COVID-19 cases are currently surging. While early detection and isolation were identified as important non-pharmaceutical intervention (NPI) measures for containing the disease spread, this may not be pragmatically implementable in developing countries primarily due to social and economic reasons (i.e. poor education, less public awareness, massive unemployment). To shed light on COVID-19 transmission dynamics and impacts of NPI scenarios – e.g. social distancing, this study conducted emerging pattern analysis using the space-time scan statistic at district and thana (i.e. a sub-district or ‘upazila’ with at least one police station) levels in Bangladesh and its capital – Dhaka city, respectively. We found that the central and south eastern regions in Bangladesh are currently exhibiting a high risk of COVID-19 transmission. Dhaka megacity remains as the highest risk “active” cluster since early April. The space-time progression of COVID-19 infection, when validated against the chronicle of government press releases and newspaper reports, suggests that Bangladesh have experienced a community level transmission at the early phase (i.e., March, 2020) primarily introduced by Bangladeshi citizens returning from coronavirus-affected countries in the Europe and the Middle East. A linkage is evident between the violation of NPIs and post-incubation period emergence of new clusters with elevated exposure risk around Bangladesh. This study provides novel insights into the space-time patterns of COVID-19 transmission dynamics and recommends pragmatic NPI implementation for reducing disease transmission and minimizing impacts in a resource-scarce country with Bangladesh as a case-study example.

## 1. Introduction

The novel coronavirus (COVID-19) infection, caused by severe acute respiratory syndrome coronavirus 2 (SARS-CoV-2) (Peeri et al., 2020), first emerged in December of 2019 in Wuhan city, China (Acter et al., 2020; Li et al., 2020). As the June 16^th^ 2020, there are more than 7.82 million confirmed cases and larger than 432,000 deaths worldwide (Dong, Du, & Gardner, 2020). Countries around the world are working to “flatten the curve” of this pandemic and many have seen a downward trend in the number of daily new cases such as Italy and Spain. However, this pandemic is still strongly present with a daily upward curve in other countries including Bangladesh. As the mid-June, Bangladesh has documented 94,481 confirmed cases in the country, with 36,264 recoveries and 1,262 deaths. The daily reports by the Institute of Epidemiology, Disease Control & Research (IEDCR) suggests that, Bangladesh detected its first three COVID-19 cases on March 8, 2020, when the number of confirmed cases already surpassed 100,000 worldwide (WHO statement, 2020). While substantial knowledge has been accumulating on the transmission dynamics of this pandemic in countries with early-state outbreak (e.g. China, Italy, United States) (Desjardins, Hohl, & Delmelle, 2020; Giuliani, Dickson, Espa, & Santi, 2020; Tang et al., 2020), relatively little is currently known of the spatio-temporal outbreak dynamics of this pandemic in Bangladesh where it’s still unfolding.

While public and private sectors in Bangladesh keep fighting to cope with the ramifications of this deadly pandemic, the scientific communities are contributing toward improving existing understanding on various aspects of this disease (Bhuiyan, Sakib, Pakpour, Griffiths, & Mamun, 2020; Chowdhury et al., 2020; Howlader, Khan, & Islam, 2020; Monjur & Hassan, 2020; Wadood et al., 2020). Among very few studies on COVID-19 spread in Bangladesh, Kalam & Hussain (2020) used analytical models (e.g. logistic) for analyzing an early growth dynamics of disease transmission. Their findings suggest that early growth of COVID-19 cases follows an exponential pattern with an estimated 5.16 days doubling period. Ahmed & Rahman (2020) reported that, COVID-19 trend in Bangladesh showed deviation from epidemiological model and identified potential factors responsible for this deviation. Overall, Bangladesh’s COVID-19 characteristic patterns reflect the followings: an inadequate testing and treatment capacity, insufficient supply of personal protection equipment, poor health services, lack of public awareness, large-scale violation of social distancing order (Figure 1 & 2), a large vulnerable population (e.g. Rohingya refugees), community-level transmission, and dire economic impacts (Islam, Talukder, Siddiqui, & Islam, 2020). While existing research and unpublished reports highlight these important aspects of the COVID-19 outbreak in Bangladesh, knowledge is largely missing on the geographic perspectives of the transmission dynamics and intervention scenarios in the country.

**Figure 1.**
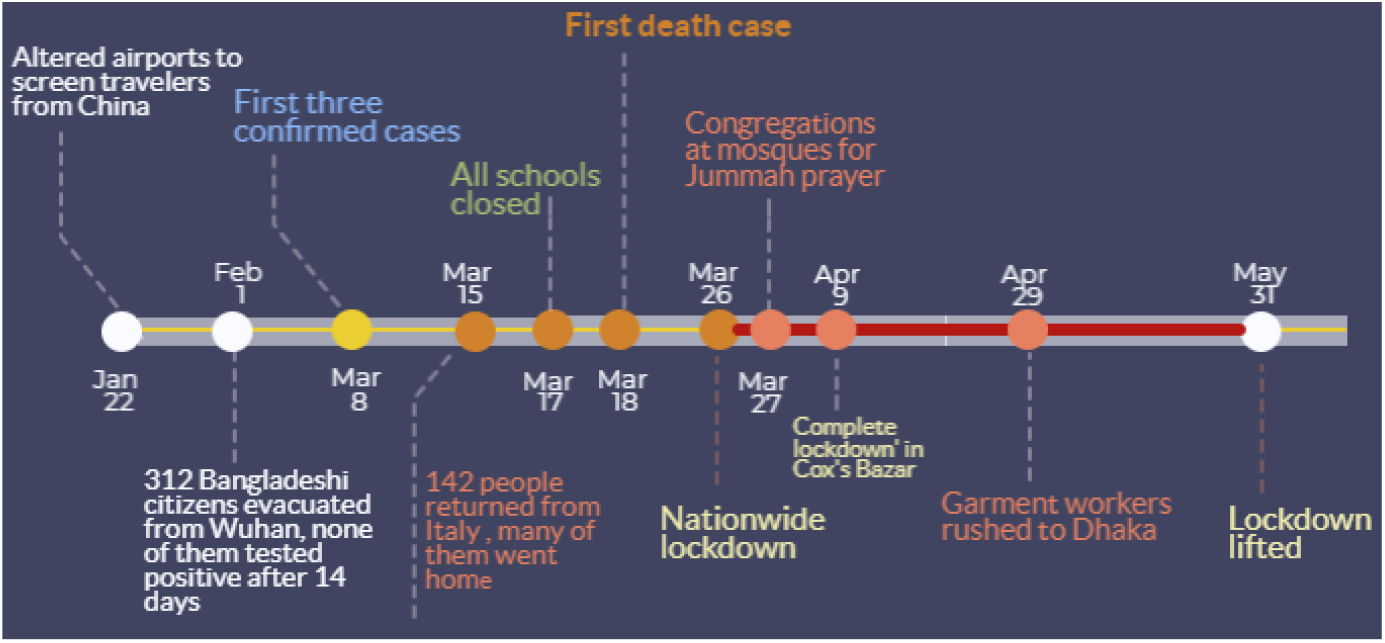
Major COVID-19 related events in Bangladesh and NPI measures by the government

**Figure 2.**
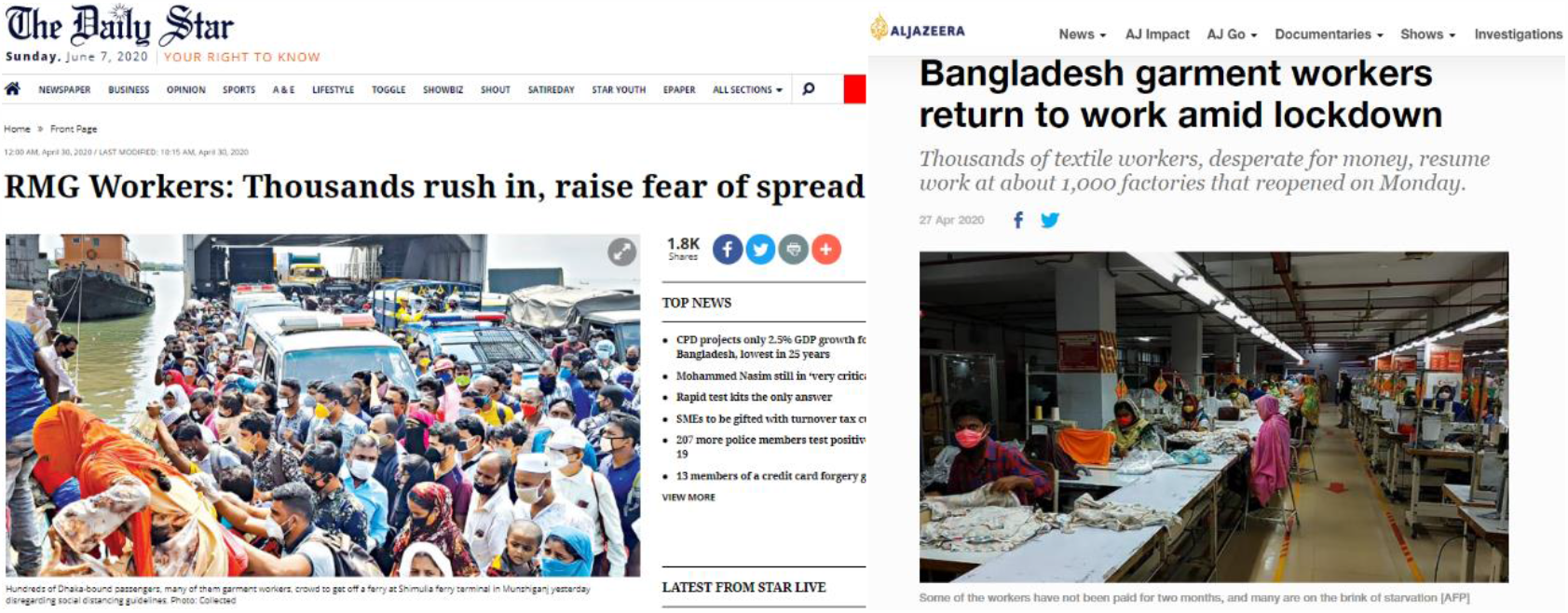
Snapshots of print media reports between April 27 and April 30 when a large number of people in Bangladesh congregated outside amidst nationwide lockdown

Spatial, temporal, and spatio-temporal statistics are essential approaches for identifying transmission dynamics of COVID-19 which can enable evaluating ongoing efforts as well as inform new, innovative solutions for containing the disease (Franch-pardo et al., 2020; Xiong et al., 2020). Existing literature shed light on the spatiotemporal dynamics of COVID-19, that helped identify the extent and impact of the pandemic as well as intervention strategies in different parts of the world including China (Guan et al., 2020; Li et al., 2020; Su et al., 2020), Italy (Gatto et al., 2020), Spain (Orea & Áacute;lvarez, 2020), (Ferreira et al., 2020), and the United States (Desjardins et al., 2020; Brazil Hohl, Delmelle, & Desjardins, 2020; Mollalo, Vahedi, & Rivera, 2020). In this paper, we leveraged a prospective space-time scan statistic approach (Kulldorff et al., 2005) to detect currently “active” and emerging clusters of COVID-19 outbreak in Bangladesh. For that, we utilized spatio-temporally aggregated daily confirmed cases made available by the IEDCR. Additionally, we conducted tracking analysis using confirmed cases of a cumulative 1-week progression (i.e. to account for incubation period) for mapping spatial propagation of the disease outbreak to understand COVID-19 transmission dynamics in both space and time. Finally, we identified potential time-lagged (i.e. characterizing incubation period) links between the space-time outbreak and NPI scenarios based on available governmental and newspaper reports.

Specifically, this study has following contributions to the understanding of COVID-19 transmission dynamics relative to NPI efforts in the example of Bangladesh:

1. Timely detection and evaluation of active space-time clusters of COVID-19 infections that currently pose threat to public health;
2. Track space-time propagation of previously existed clusters for an understanding of ‘where’ and ‘when’ infections spread;
3. Identify linkages between the retrospective emerging patterns of COVID-19 spread and time-lagged NPI scenarios.

## 2. Materials and Methods

### 2.1 COVID-19 Daily Reports – Bangladesh and Dhaka megacity

We used freely available COVID-19 daily reports from the IEDCR website (https://www.iedcr.gov.bd/) that contain cumulative counts of confirmed cases over the past 24 hours nationwide. The daily confirmed cases had been on the rise (mid-June 2020) in Bangladesh, particularly in Dhaka megacity. Although district-level reported cases were made available since March 4 by IEDCR, currently location-based (i.e. place of the patient’s resident) daily counts are only available from April 4 Bangladesh and April 7 for Dhaka. Figure 3 shows cumulative counts of confirmed, recovered, and death cases at the country-level.

**Figure 3.**
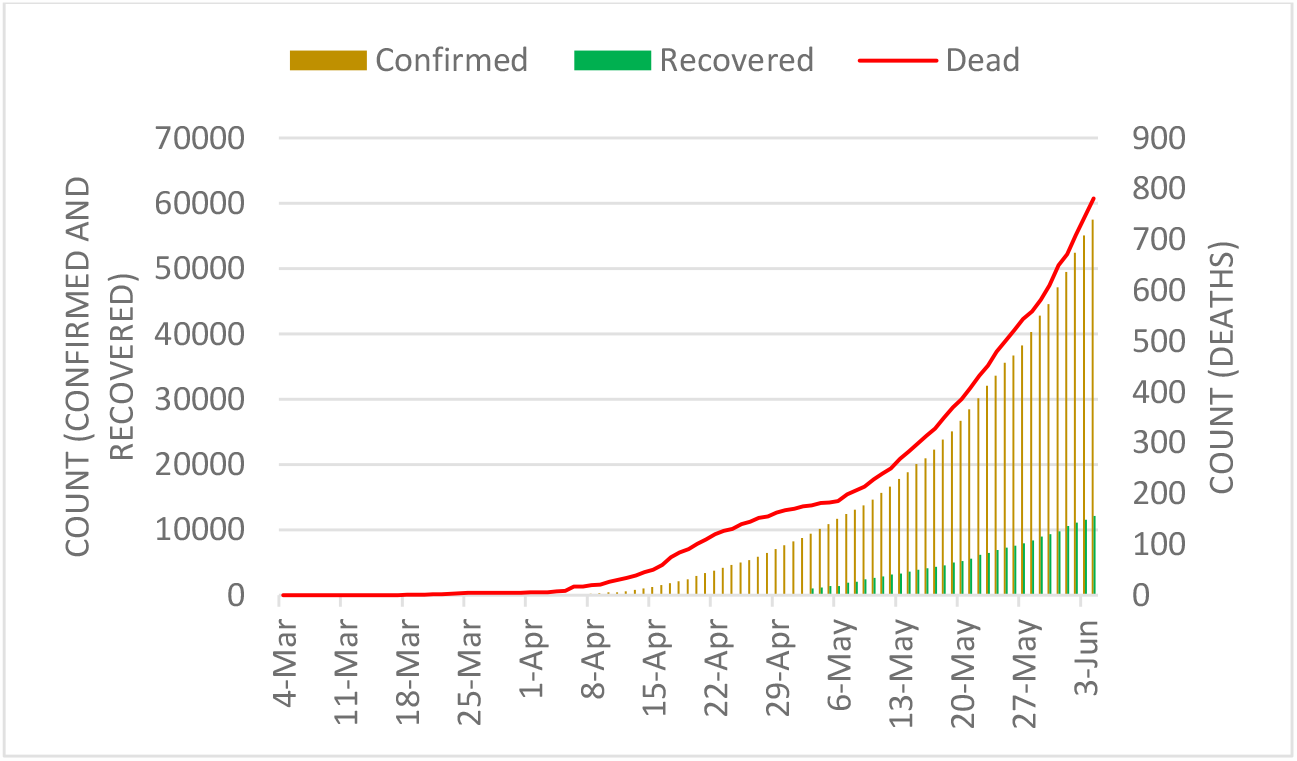
Cumulative count of COVID-19 cases in Bangladesh

Our emerging cluster analyses required counts of new cases per temporal unit (e.g., day) at different geographic locations – i.e. districts and thanas. Therefore, we subtracted daily cumulative counts from the previous day’s count to obtain the present day’s new cases for the entire available timeline. Figure 4 shows daily counts of newly confirmed, recovered, and death cases over the past 24 hours at the country-level (i.e., sum of all districts per day).

**Figure 4.**
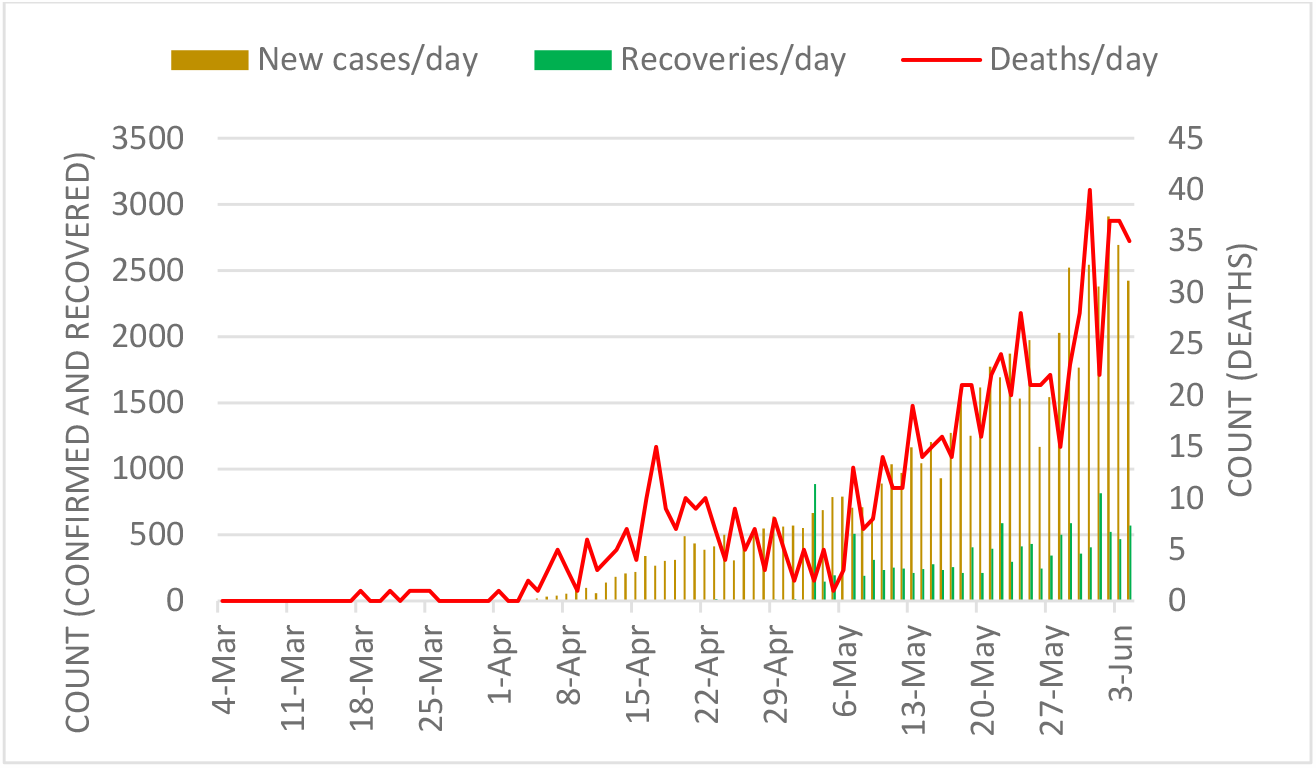
Daily new cases of COVID-19 cases in Bangladesh

### 2.2 LandScan Population Data

Our analysis required the specification of population size over space in each spatial unit. For that, we used the LandScan™ Population dataset with ∼1km spatial resolution (Dobson, Bright, Coleman, Durfee, & Worley, 2000). Currently available 2018 LandScan dataset of Bangladesh was used to extract total population count at the thana level for Dhaka city. Note that, our district-level analysis used the population count that was inherently present with the COVID-19 dataset for Bangladesh provided by IEDCR’s GIS dashboard (IEDCR, 2020).

### 2.3 Emerging space-time cluster analysis

We detected currently emerging space-time clusters of COVID-19 cases in Bangladesh using the prospective space-time scan statistic (Kulldorff, 2001; Kulldorff et al., 1998). For our analyses, we used SaTScan™ (Kulldorff, 2018) (version. 9.6). The SaTScan statistic has been regularly used in epidemiology to identify significant clusters of diseases. Using different models (e.g. Poisson, Bernoulli) it detects and maps unexpected space-time clustering given baseline conditions (e.g. population size). This statistic is defined by a cylinder-shaped window with a round spatial base and with height analogous to time. The base of the cylinder is centered on centroids of the geographic unit (e.g. district) throughout the study area (i.e. Bangladesh). The radius of the base varies continuously in size until a maximum upper bound is reached. The height denotes time interval of <= 50% of the study period, as well as the study period as a whole. The cylindrical window is moved in space and time to cover each possible geographic location and size, as well as each possible time interval.

The prospective space-time scan statistic detects active clusters of COVID-19 infections that are still being found at the end of the study period. Table 1 shows input parameters used in our prospective analyses. We used a discrete Poisson-based probability model on the assumption that the COVID-19 cases follow a Poisson distribution according to the at-risk population in Bangladesh. The model also assumes the population size to be static in an area over a time period.

**Table 1.** Parameter setup for the prospective Poisson space-time scan statistic

| **Parameters used in prospective analysis** |  |
| --- | --- |
| **Probability model** | Discrete Poisson |
| **Scan for areas** | High or low rates |
| **Time aggregation units** | 1 |
| **Spatial window** | 10% of the population at risk |
| **Spatial window shape** | Circular |
| **Maximum temporal cluster size** | 50% of the study period |
| **Minimum temporal cluster size** | 2 units |
| **Minimum number of cases for high rate clusters** | 3 (5 for Dhaka city) |
| **p-value** | Default |
| **Maximum Monte Carlo replications** | 999 |
| **Spatial and temporal trend adjustments** | None |

The null hypothesis H_0_ stipulates that the model represents a diverse Poisson process of contracting COVID-19 with an intensity μ (proportional to the at-risk population), whereas the alternative hypothesis H_A_ supports that the number of observed cases is greater than the number of expected cases. The following equation calculates an expected number of COVID-19 cases (μ) (Desjardins et al., 2020; Owusu et al., 2019):

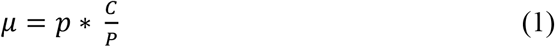

where, p is the population in a geographic area (e.g. district or thana); C and P are the total COVID-19 cases and the total estimated population in Bangladesh, respectively.

For each cylinder, the number of infections inside and outside the cylinder, as well as the expected number of cases reflecting the population at risk and relevant covariates are used to calculate the likelihood for each cylinder. The most likely cluster has a cylinder with the maximum likelihood and more than the expected number of cases. Equation 2 was used for calculating maximum likelihood ratio that identified scanning windows with elevated risk for COVID-19 (Kulldorff, 2001).

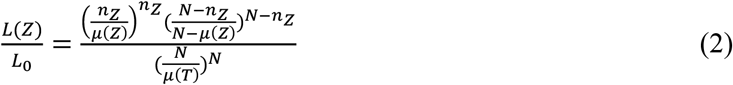

where, L(Z) the likelihood function for Cylinder Z, and L_0_ the likelihood function for H_0_; n_Z_ the number of COVID-19 cases in a cylinder; μ(Z) the number of expected cases in Cylinder Z; N being the total number of observed cases for the entire study areas – Bangladesh or Dhaka city, across all time periods; and μ(T) the total number of expected cases in the study area across all time periods. Risk elevates for the cylinder when it has a likelihood ratio greater than 1.

Multiple geographic units can belong to a significant space-time cluster, which assumes that the relative risk (RR) of COVID-19 is homogeneous across a cluster. To avoid that assumption, we derived relative risk for each geographic unit – district and thana – that lies within a cluster. The RR is calculated using Equation 3 (Desjardins et al., 2018; Owusu et al., 2019):

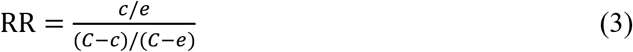

where, c is the total number of COVID-19 cases in a district, e is the total number of expected cases in a district, and C is the total number of observed cases in Bangladesh, for example. RR is thus an estimated risk within a location divided by the risk everywhere else. Clusters also have RR which is calculated using same way as in Equation 3. For example, a RR value of 5.5 for a location or a cluster suggests that the population within this location or cluster is 5.5 times more likely to be exposed to COVID-19 infections compared outside of that location or cluster.

In the following section, we present active and emerging clusters of COVID-19 cases in the Dhaka city and all 64 districts of Bangladesh. To elucidate the space-time transmission dynamics of COVID-19 cases in Bangladesh relative to NPI scenarios, first, we divided current timeline of COVID-19 cases into two groups: April 4 to April 30 and April 4 to June 5. April 30 was used as a cut-off date as it signifies one of the major breaks in social distancing over the past two weeks (Figure 2), which may have led to the emergence of new active clusters. Second, for an improved understanding of the space-time propagation of relative risk across districts in Bangladesh, we detected emerging clusters using a cumulative 1-week prospective scanning approach. This enabled considering a roughly 1 to 2-week incubation period for locating dissipating risk and newly emerged high-risk areas along the timeline. Finally, we analyzed the linkages between these clusters and known NIP scenarios in space and time.

## 3. Results

### 3.1 Thana-level emerging clusters in Dhaka megacity

Being the capital and largest industrial and economical hub of the country, Dhaka experienced the inception of the COVID-19 outbreak in Bangladesh. Although the Bangladesh government initiated intervention measures in mid-March (see Figure 1), it was not until March 26 when the entire nationwide lockdown was put in place and extended until May 31, 2020. Meanwhile, other intervention measures were neglected by the authorities and general public (Kamruzzaman, 2020) which may have resulted in the transmission of COVID-19 across and beyond Dhaka city.

We found eleven statistically significant emerging space-time clusters of COVID-19 at the thana level in Dhaka megacity from April 7 to June 15, 2020. Table 2 shows that among these clusters there were twenty-three thanas with a higher relative risk (RR>1), where there were more observed than expected coronavirus cases over the timeline based on the parameters set in Table 1. These high RR cluster areas vary in size and magnitude of risk. Lower RR (< 1) areas are characterized by higher expected than observed cases. The relationship between population density and relative risk suggests that all locations in Dhaka have not responded evenly to the outbreak. For example, the emerging cluster with the highest RR (11.94) is found in Mirpur with a relatively moderate population size (Figure 5 & Appendix A1). Other areas within Pallabi and Rupnagar thana that are relatively more populated and lower to medium-income were experiencing a relatively high level of RR (4.42). The northern most (i.e. Uttara thana) areas in Dhaka city were exhibiting smaller radius of emerging hotspot but with higher magnitudes. The onset of these “active” clusters took place between early May and mid-June. However, clusters that emerged early (May 12 – June 15) were exhibiting lower risk, whereas relatively younger clusters (May 30 – June 15) demonstrated higher risk of outbreak. At the same time, many areas were exhibiting low to no risk by mid-June, which were high-risk areas over the early April to mid-May period. A weekly or bi-weekly prospective scanning will reveal the space-time propagation of COVID-19 hotspots in Dhaka city. Since our primary focus is on elucidating district level transmission dynamics, in the next section we have demonstrated space-time emerging patterns and propagation of COVID-19 for the entire country.

**Table 2.** Emerging space-time clusters of COVID-19 in Dhaka city from April 7 - June 15, 2020

| Cluster | Duration (days) | No. of<br>Thanas | *p* | Observed | Expected | Relative<br>Risk<br>(RR) | No. of<br>thanas<br>RR > 1 |
| --- | --- | --- | --- | --- | --- | --- | --- |
| **1** | 30-May - 15-Jun | 1 | <0.001 | 1289 | 107.98 | **11.94** | 1 (Mirpur) |
| **2** | 30-May - 15-Jun | 6 | <0.001 | 1304 | 294.99 | **4.42** | 6 |
| **3** | 30-May - 15-Jun | 5 | <0.001 | 1142 | 332.11 | **3.44** | 5 |
| **4** | 30-May - 15-Jun | 1 | <0.001 | 506 | 106.85 | **4.74** | 1 (Uttara) |
| **5** | 30-May - 15-Jun | 2 | <0.001 | 539 | 163.76 | **3.29** | 2 |
| **6** | 12-May - 15-Jun | 3 | <0.001 | 177 | 664.8 | 0.27 | 0 |
| **7** | 30-May - 15-Jun | 5 | <0.001 | 648 | 300.39 | **2.16** | 5 |
| **8** | 30-May - 15-Jun | 2 | <0.001 | 143 | 38.5 | **3.71** | 2 |
| **9** | 12-May - 15-Jun | 1 | <0.001 | 152 | 305.51 | 0.5 | 0 |
| **10** | 12-May - 15-Jun | 5 | <0.001 | 262 | 428.88 | 0.61 | 1 |
| **11** | 12-May - 15-Jun | 1 | <0.001 | 97 | 203.16 | 0.48 | 0 |

*Note:* RR > 1.0 and the highest RR are highlighted in bold and red colored texts, respectively.

**Figure 5.**
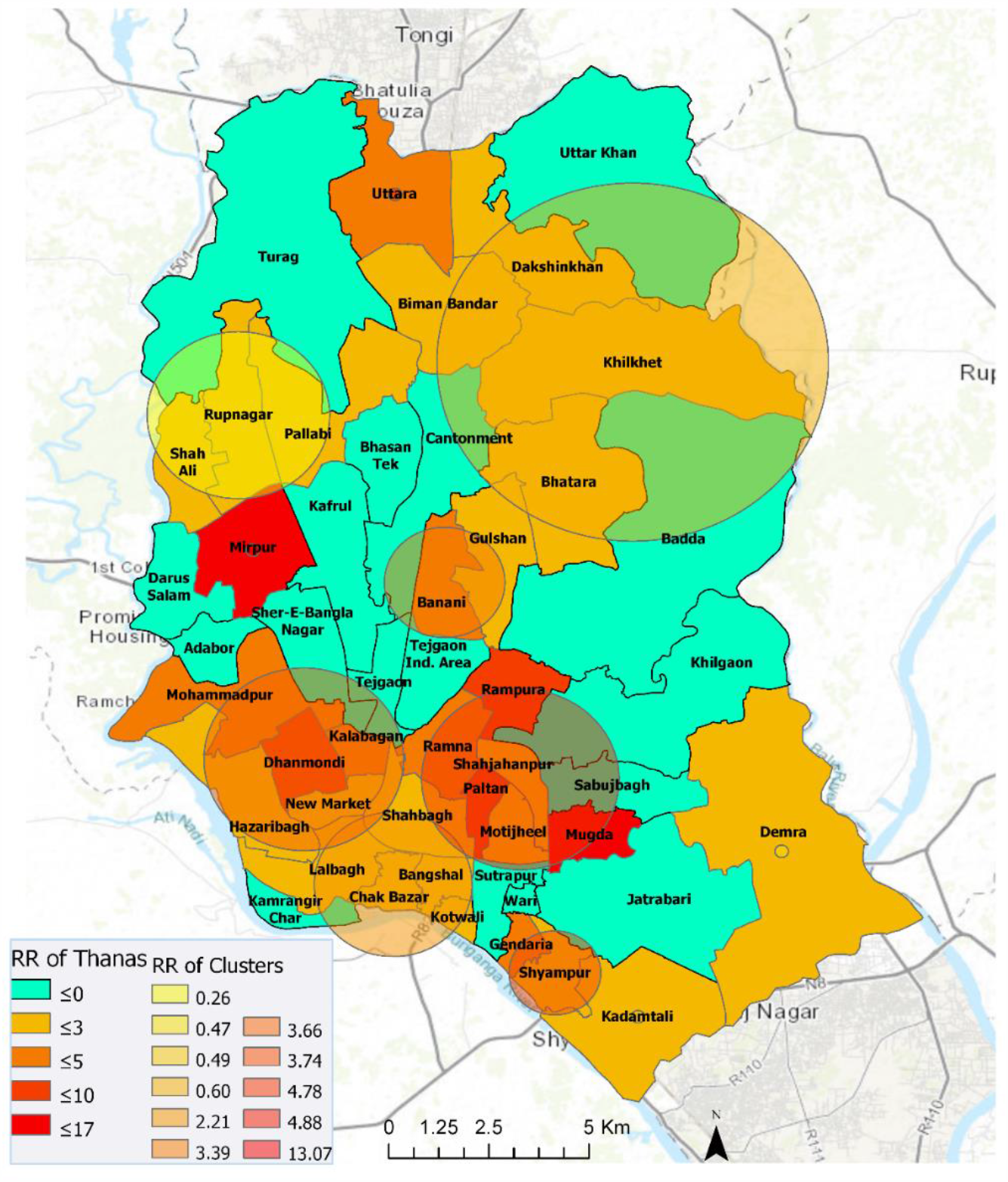
Emerging space-time clusters at the thana-level in Dhaka city (April 7 - June 15, 2020)

### 3.2 District-level emerging clusters - April 4 – June 5, 2020

As mentioned previously, travel to and from Dhaka city, before and during the nationwide lockdown, may have propagated a community-level transmission in other parts of the country. Hence, in the next sections, we show results from district-level analyses for two different periods.

#### 3.2.1 April 4 – April 30, 2020

Ten statistically significant emerging space-time clusters were detected in Bangladesh between April 4 and April 30. Table 3 shows characteristics of these clusters. There were two high-risk clusters with RR > 1 (i.e., more observed cases than expected) and eight low-risk clusters with RR < 1 (i.e., more expected than observed) across a total of 46 districts with at least one confirmed COVID-19 case. During this period, 20 districts out of total 64 districts in Bangladesh exhibited no relative risk of exposure (RR = 0) to the COVID-19 infection (Figure 6). Cluster 1, containing Dhaka (metropolitan area), Narayanganj, and Munshiganj, had a RR of 25.44, which suggests that the population within this cluster was a record 25.44 times more likely to be exposed to COVID-19 compared to other clusters. Cluster 9 contains only one district – Kishoreganj, with a RR of 2.08. The other eight clusters had RR less than 1, suggesting that population within these cluster locations were exposed to a very low-risk of coronavirus infection. Cluster 10 that included two districts – Narsingdi and Brahamanbaria – exhibited smaller and a relatively delayed time period (April 22 – April 30) than other clusters. Note that, this low-risk exposure in Brahamanbaria elevated to a higher-risk exposure in the following week (Figure 7 & 9), which altogether can be linked to a public event in the previous week (on April 18) that violated government-imposed social distancing order (Table 5). Figure 6 visualizes locations of high and low RR districts within these ten clusters.

**Table 3.** Emerging space-time clusters of COVID-19 at the district level between April 4 and April 30, 2020

| Cluster | Duration (days) | No. of Districts | *p* | Observed | Expected | Relative Risk | No. of districts<br>RR > 1 |
| --- | --- | --- | --- | --- | --- | --- | --- |
| **1** | 18-Apr - 30-Apr | 3 | <0.001 | 3866 | 310.1932 | **25.44** | 3 |
| **2** | 18-Apr - 30-Apr | 6 | <0.001 | 56 | 349.8174 | 0.15 | 0 |
| **3** | 18-Apr - 30-Apr | 6 | <0.001 | 68 | 332.197 | 0.20 | 0 |
| **4** | 18-Apr - 30-Apr | 6 | <0.001 | 48 | 288.4021 | 0.16 | 0 |
| **5** | 18-Apr - 30-Apr | 4 | <0.001 | 73 | 267.9812 | 0.27 | 0 |
| **6** | 18-Apr - 30-Apr | 8 | <0.001 | 100 | 281.342 | 0.35 | 0 |
| **7** | 18-Apr - 30-Apr | 6 | <0.001 | 126 | 318.2578 | 0.39 | 0 |
| **8** | 18-Apr - 30-Apr | 4 | <0.001 | 45 | 126.2793 | 0.35 | 0 |
| **9** | 18-Apr - 30-Apr | 1 | <0.001 | 147 | 71.30506 | **2.08** | 1 |
| **10** | 22-Apr - 30-Apr | 2 | <0.001 | 34 | 89.81475 | 0.38 | 0 |

*Note:* RR > 1.0 and the highest RR are highlighted in bold and red colored texts, respectively.

**Fig 6.**
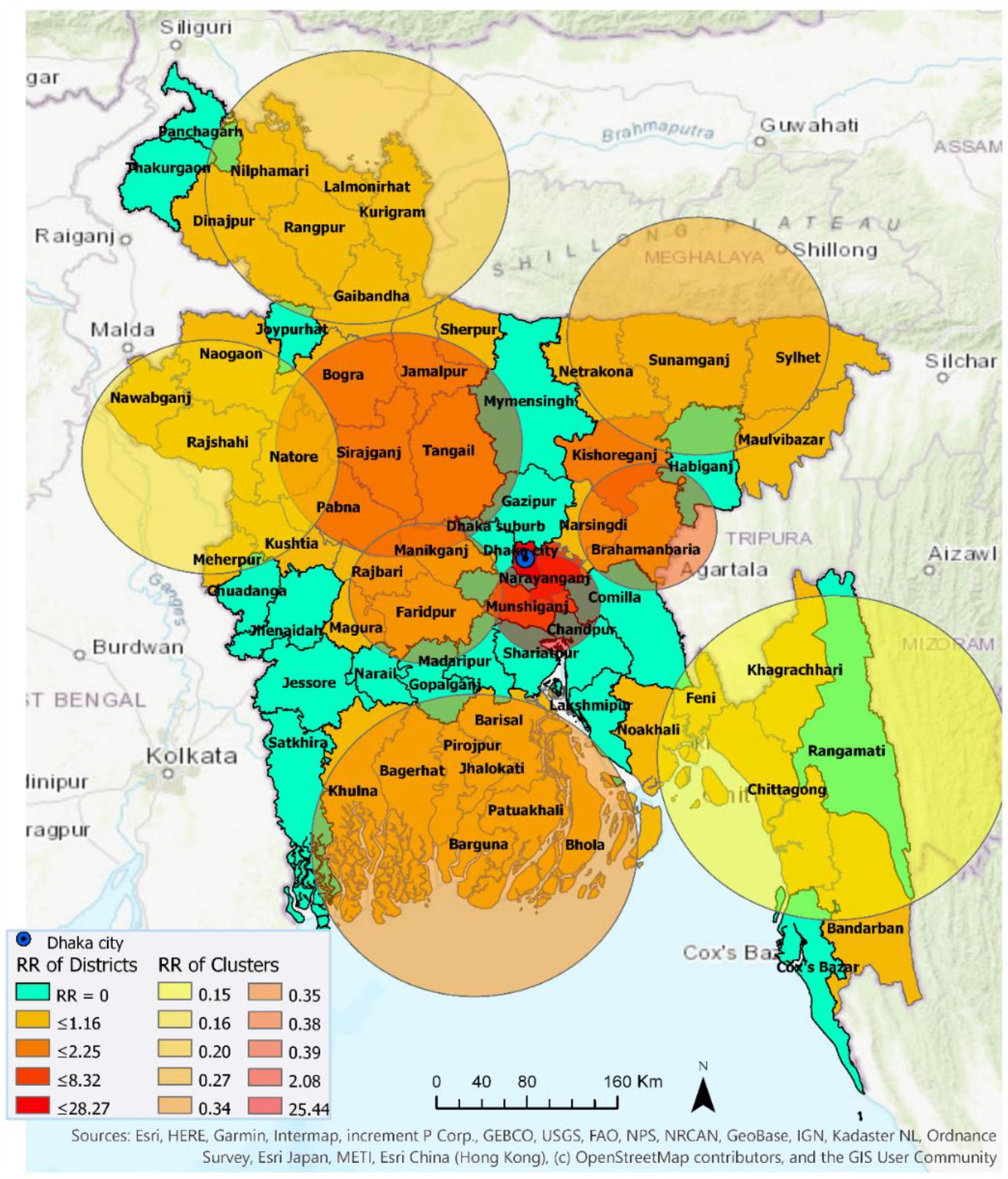
Emerging space-time clusters at the district level between April 4 and April 30, 2020

**Figure 7.**
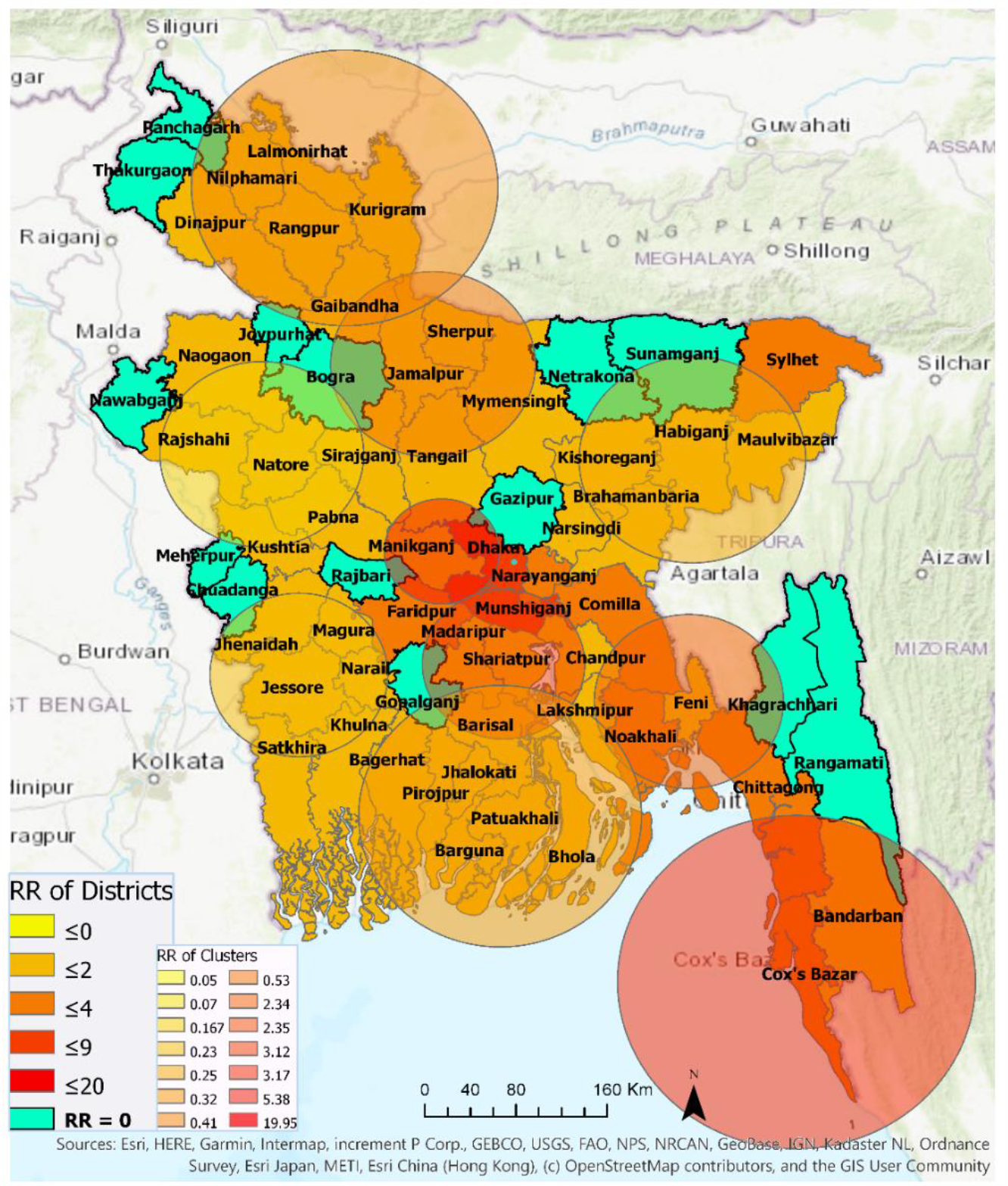
Emerging and non-emerging space-time clusters at the district level between April 4 and June 5, 2020

#### 3.2.2 April 4 – June 5, 2020

Fourteen statistically significant emerging space-time clusters were detected in Bangladesh between April 4 and June 5. Table 4 summarizes characteristics of these clusters. The number of clusters with RR > 1 increased to six. More districts (51) are included in these emerging clusters with 16 districts contained by high-risk clusters (RR >= 2.35) and the other 35 districts are considered low-risk clusters. The most likely high-risk cluster – i.e. Cluster 1, with the highest RR of 19.95 contains only one location – Dhaka metropolitan area (RR = 19.946201). Suburban Dhaka (RR = 9.20) belongs to the next high cluster, i.e. Cluster 15 (RR = 5.38). This cluster contains two neighboring districts: Faridpur (RR = 3.03) and Manikganj (RR = 0.78). The third high cluster (Cluster 2) with RR of 3.17 contains three southeastern districts: Chittagong (RR = 3.21), Cox’s Bazar (RR = 3.14), Bandarban (RR = 0.71). The fourth high-risk cluster (Cluster 4) with RR of 3.12 neighbors the first and second high clusters, and contains five districts: Munshiganj (RR = 6.09), Narayanganj (RR = 4.16), Madaripur (RR = 1.85), Shariatpur (RR = 1.07), and Chandpur (RR = 1.02). The fifth high-risk cluster (Cluster 14) with RR of 2.35 contains only one district, which is the most northeastern district of Bangladesh - Sylhet. Figure 7 shows that most of the northwestern and southwestern districts belong to low-risk clusters. There are 13 non-emerging COVID-19 risk districts in Bangladesh at the time of this analysis, they are Bogra, Chuadanga, Gazipur, Gopalganj, Joypurhat, Meherpur, Nawabganj, Netrakona, Panchagarh, Rajbari, Rangamati, Sunamganj, and Thakurgaon.

**Table 4.** Emerging space-time clusters at the district level, between April 4 and June 5, 2020

| Cluster | Duration (days) | No. of Districts | *p* | Observed | Expected | Relative Risk (RR) | No. of districts<br>RR > 1 |
| --- | --- | --- | --- | --- | --- | --- | --- |
| **1** | 6-May - 5-Jun | 1 | <0.001 | 14189 | 1069.302 | **19.95** | 1 |
| **2** | 13-May - 5-Jun | 3 | <0.001 | 3457 | 1160.286 | **3.17** | 3 |
| **3** | 6-May - 5-Jun | 5 | <0.001 | 287 | 1665.789 | 0.17 | 0 |
| **4** | 22-May - 5-Jun | 5 | <0.001 | 1907 | 630.8265 | **3.12** | 5 |
| **5** | 30-May - 5-Jun | 3 | <0.001 | 978 | 185.429 | **5.38** | 3 |
| **6** | 6-May - 5-Jun | 6 | <0.001 | 342 | 1424.87 | 0.23 | 0 |
| **7** | 7-May - 5-Jun | 4 | <0.001 | 357 | 1399.596 | 0.25 | 0 |
| **8** | 8-May - 5-Jun | 8 | <0.001 | 499 | 1520.132 | 0.32 | 0 |
| **9** | 22-May - 5-Jun | 3 | <0.001 | 1595 | 696.3971 | **2.35** | 1 |
| **10** | 6-May - 5-Jun | 6 | <0.001 | 790 | 1877.635 | 0.41 | 0 |
| **11** | 6-May - 5-Jun | 1 | <0.001 | 25 | 488.2511 | 0.05 | 0 |
| **12** | 6-May - 5-Jun | 1 | <0.001 | 24 | 320.769 | 0.07 | 0 |
| **13** | 6-May - 5-Jun | 4 | <0.001 | 846 | 1553.26 | 0.53 | 0 |
| **14** | 22-May - 5-Jun | 1 | <0.001 | 628 | 269.7846 | **2.34** | 3 |

*Note:* RR > 1.0 and the highest RR are highlighted in bold and red colored texts, respectively.

Table 4 shows that, emerging space-time clusters manifest varying onset time and duration. Among the high-risk clusters, Cluster 1 that contains Dhaka megacity (i.e. most likely cluster) emerged as the major hotspot on May 6 and remained so at the time of this analysis. Cluster 2 is the second-longest (i.e. 24 days or > 3 weeks) active high-risk cluster that is primarily characterized by industrial (e.g. seaports) and tourist areas (e.g. sea beaches, hill resorts). Other three relatively high risk and spatially contiguous clusters – Clusters 4, 9, and 14 – emerged on the same day on May 22. Cluster 5 is the most recently emerged (on May 30) and second high-risk cluster that contains Dhaka’s suburb regions with industries and garment factories.

### 3.3 Progression of relative risk of COVID-19

#### 3.3.1 Comparison between early and later phase (pre- and post-April 30)

Figure 8 highlights the changes in relative risk (RR) of COVID-19 in Bangladesh at the district level between two emerging periods: April 4 – April 30 and a longer period of April 4 – June 5. In essence, this temporal change in RR indicates space-time patterns of transmission, most likely responding to spatiotemporally variable levels of non-pharmaceutical intervention measures practiced by people. Out of the sixty-four districts in Bangladesh, seven districts manifested RR = 0 over the two periods. Thus, they are categorized as “non-emerging” COVID-19 regions, although it should be noted that all of these seven districts (Panchagarh, Thakurgaon, Chuadanga, Joypurhat, Gazipur, Gopalganj, and Rangamati) have experienced COVID-19 outbreak (i.e. > 100 cases in each of these districts) and some of them became emerging clusters (with elevated RR) at some point in time when scanned over a shorter temporal window (see Figure 9). Besides, different parameterization for the space-time scanning, including the duration of emerging clusters and minimum number of confirmed cases, may provide slightly different patterns.

**Figure 8.**
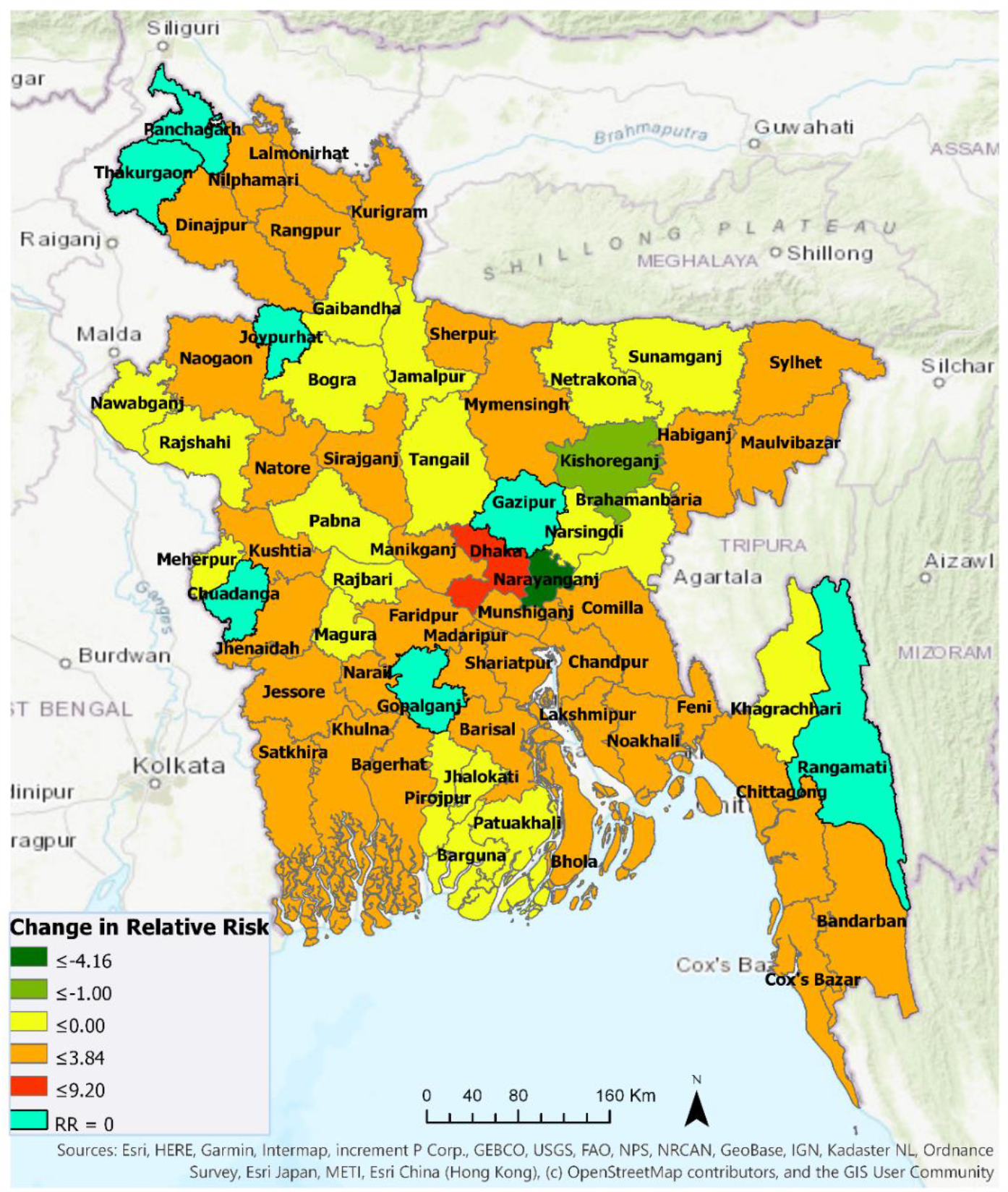
Changes in relative risk of COVID-19, between two emerging periods (April 4 – April 30 and April 4 – June 5) in Bangladesh

**Figure 9.**
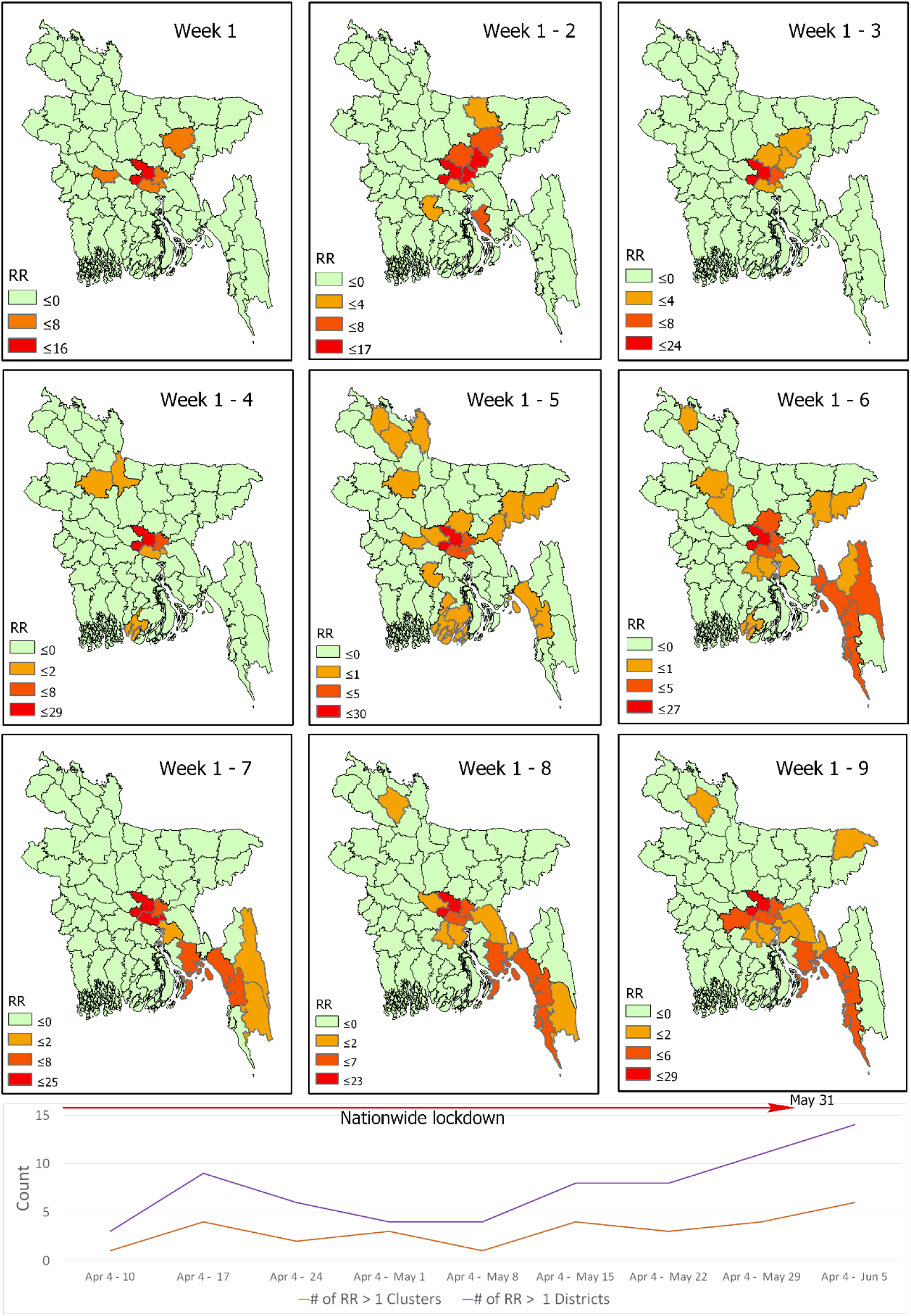
Weekly temporal pattern of space-time propagation of relative risk of COVID-19 in Bangladesh

The suburban region of Dhaka, which is characterized high-density working-class population, emerged as a new high-risk cluster with RR of 9.2 during May 1-June 5, likely responding to a large-scale violation of social distancing in the late April. Although Dhaka metropolitan area experienced a reduced level of risk (-8.3 change in RR), it remained the epicenter of outbreak with the highest risk level (RR =19.95) over the same period. Similarly, the relative risk in Narayanganj and Kishoreganj districts decreased as well, although Narayanganj remained one of the high-risk districts. The areas that manifested > 3.00 increase in RR over this period were: Munshiganj, Chittagong, and Cox’s Bazar. Districts with > 2.00 increase in RR were: Faridpur, Comilla, and Noakhali, followed by Madaripur, Shariatpur, Chandpur, and Feni where RR increased by 1. The relative risk of COVID-19 increased all over the country after April 30 in Figure 8. This essentially portrays a local community-level transmission all around Bangladesh. However, it is noticeable that the relatively higher risk areas are in the central and southeastern districts. Whereas, some southern districts progressed toward reduced risk of an outbreak. Hilly district Khagrachari turned into “non-emerging” areas with RR dropped to 0, whereas Bandarban experienced slightly elevated risk with +28 cases. This relatively lower to no risk pattern in the hilly districts could be related to lower population density and restricted movement patterns due to inaccessible locations characterised by high altitude than majority of Bangladesh lands.

#### 3.3.2 Tracking of changing space-time clusters

Figure 9 shows space-time progression of COVID-19 disease in Bangladesh at weekly intervals. Based on the available data, nine weekly intervals were used for space-time scanning between April 4 and June 5. Most of the districts had RR = 0 in week 1, except for Dhaka (mostly metropolitan area) and its industrial belts in Narayanganj, Munshiganj, Kishoreganj, and Rajbari. It should be noted that many Bangladeshi expatriates from Europe and Middle-East returned in Dhaka and subsequently returned to their home districts by early April. Besides Dhaka and its contiguous neighbors being the epicenter of the COVID-19 outbreak, the emergence of Kishoreganj and Rajbari as high-risk areas at this relatively early-phase could be related to the presence of returnees from foreign countries and the epicenter itself. Week 2 shows expansion as well as an increased level of risk toward north and east of Dhaka to include Gazipur and Narsingdi. Additionally, Netrakona, Gopalganj, and Lakshmipur emerged as high-risk areas. Week 3 is characterized by lower-risk areas, which may well just reflect the missing data issue. Later weeks show how COVID-19 transmitted to more areas with elevated risk. By week 5 (May 2 – 8), COVID-19 hotspots reached northeastern Bangladesh. In the following weeks (May 9 – June 5), southern coastal and hilly districts in southeastern Bangladesh emerged as hot-spots. Although the Bangladesh government lifted nationwide lockdown on May 31 for economic reasons, Figure 9 clearly shows the existence of active hotspots particularly in the industrial belts of Dhaka and Chittagong.

With an understating of the weekly progression of emerging clusters, we can begin to link these emergences with intervention scenarios in accordance with the incubation period which can be up to 2 weeks. Table 5 shows some currently available empirical evidence that may highlight a potential link between detected space-time risk progression in Figure 9 and public events associated with social-distancing measures. An average incubation period of 2-3 weeks between the exposure and emergence events might support these linkages. However, the emergence of Rajbari as a high RR district on the week of April 4 -10 may not be directly linked to the shutdown of Daulatdia brothel. In contrast, coronavirus in Rajbari could have been transmitted by Bangladeshi workers returning from Italy or elsewhere. On the other hand, temporal duration between the emergence of COVID-19 risk in the south-eastern coastal districts and the prior tourist activity violating intervention measure in late March is longer than the usual incubation period of two weeks. However, this somewhat ‘delayed’ emergence may have resulted from a low testing rate of potential patients as well as the presence of asymptotic cases in Bangladesh. As more spatial epidemiological evidence and more local-level (e.g. sub-district) data become available a large-scale contact tracing of confirmed cases will likely confirm these and find additional linkages at the local and inter-regional scales.

**Table 5.**
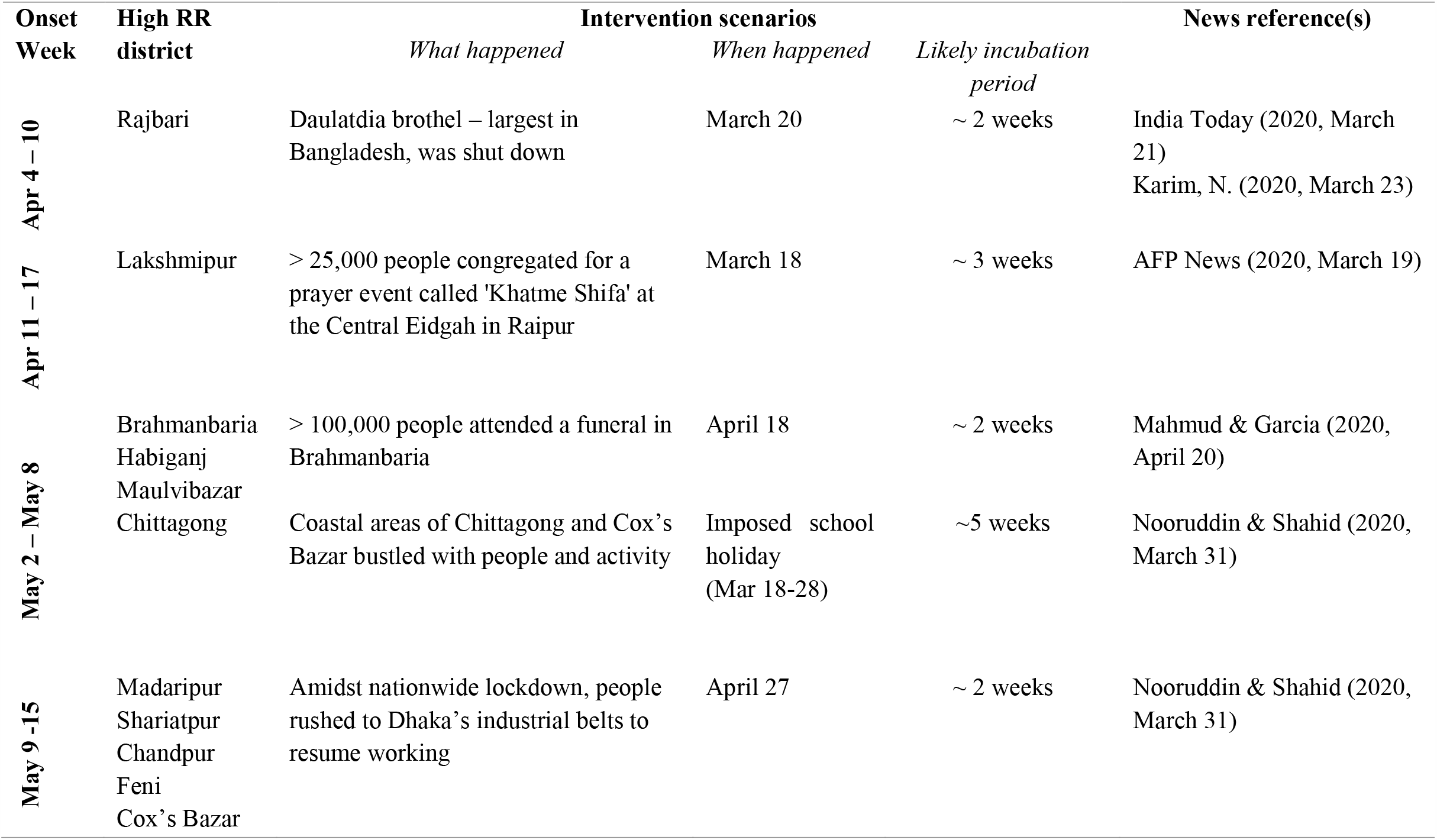
Potential linkage between space-time progression of COVID-19 risk and social-distancing violation in Bangladesh

## 4. Discussion and Conclusions

This study leveraged prospective space-time scan statistics for identifying currently active or emerging clusters of COVID-19 in Bangladesh. We conducted this analysis at the district level for Bangladesh and the thana level for the Dhaka metropolitan area - one of the most densely populated megacities in the world. This study, to the best of our knowledge, is a first attempt to showcase space-time progression of COVID-19 risk in Bangladesh. It is also the first work that utilized prospective scanning statistics to detect emerging clusters of COVID-19 in the country. As the COVID-19 cases are still on the rise as of mid-June, the prospective approach demonstrated here can be utilized for rapidly monitoring evolving space-time pattern of COVID-19 risk in Bangladesh and elsewhere. This will enable government and health officials to take appropriate and time-sensitive intervention measures and potentially prepare for future outbreaks of a highly contagious disease.

We investigated emerging space-time clusters at the thana level in Dhaka. Utilizing April 7 - June 15, 2020 of daily COVID-19 reported cases, we have identified 11 emerging clusters with 23 relatively higher magnitudes of risky thanas (Figure 5). Multiple emerging clusters are spreading all over the city with varying size and magnitude. Most densely populated and lower to medium-income locations of Dhaka city around Mirpur, Uttara, Mugda, Rampura, Mohammadpur, Ramna, Shahjahanpur, Gendaria, and Shyampur thana were experiencing a moderate to higher level of risk at the time of this analysis (i.e. mid-June). Among higher income areas, Dhanmondi and Banani were exhibiting higher risk of COVID-19 outbreak. However, many thanas were at low to no risk by June 15, such as Kamrangir Char, Bangshal, Gulshan that were higher risk areas over the early April to mid-May period. As time progresses and more data become available, using this space-time scanning measure at relatively shorter temporal interval would assist monitoring day-to-day or week-to-week transmission of previously existing and newly emerging clusters. Further evaluation with covariates such as age, sex, income, litigation measures could enable a better understanding of the rate and process of COVID-19 transmission. However, such an analysis is beyond the scope of this study.

While exploring the space-time progression of COVID-19 cases at the district level, we found that there were only two high-risk clusters in four districts in the central Bangladesh before May. This was most likely introduced by travelers coming from abroad including China, Italy, UK, Saudi Arabia, Kuwait, Bahrain, and India. However, although that is the most likely case as suggested by the chronicle of government press releases and newspaper reports, without rigorous contact tracing of COVID-19 patients in the early phase (e.g., January-February) any such assumption could not be scientifically validated. The Bangladesh government initiated measures on safe evacuations of Bangladeshi citizens from Wuhan, China, travel restrictions, and social distancing measures that may have been effective in restricting further outbreak until early March. However, very insufficient and limited testing facilities may have been contributed to that perceived scenario as well in the country. In addition, large-scale violations of the social distancing effort a few times, particularly between April 27 and April 30, contributed to the transmission, outside of Dhaka. Our analysis identified a substantial increase in the number of emerging clusters in the second phase after April 30. The onset and duration of the emerging clusters (Table 3 and 4) suggest that, Dhaka’s suburb and neighboring regions, where garment workers and other manufacturing industries workers are located, emerged as high-risk clusters exactly within 3 to 4 weeks after a social-distancing breakdown. To better understand the COVID-19 transmission dynamics, datasets on patient’s travel and contact history needs to be incorporated, which is very difficult to get at this point of time for a data sparse country.

The prospective space-time scanning results presented here suggest that, Bangladesh may well have experienced a community-level exposure to the deadly coronavirus pandemic in March. The primary transmission agents were people returning from abroad in February and March, especially from highly infected countries in Europe (e.g. Italy) and the Middle East (e.g. Saudi Arabia). Although the government started taking initiatives in March, it was belated to contain the outbreak primarily in Dhaka city. For that, early detection and effective isolation measures were necessary. In addition, the space-time progression from early April that highlights community level spread in districts was potentially facilitated by large-scale violations of social distancing order. Despite mass gatherings due to poor education as well as behavioral, and dogmatic religious practices (Figure 10), the government’s poor policy guidance and lack-luster enforcement exacerbated the COVID-19 situation. Although there are substantial differences in education and socio-economic structure, Bangladesh’s COVID-19 spread pattern could be compared to some of the developed nations such as the United States (Desjardins et al., 2020). Similar to the US cases, the Bangladesh government wasted precious time in the early phase of this pandemic for early detection and isolation, as well as missed the opportunity to rapidly organize a somewhat ‘ineffective’ public health system. Consequently, both countries experienced a rapid outbreak at the city to local levels. However, Bangladesh remains low in coronavirus-death rate per 100,000 people among top-20 countries with the highest counts of confirmed cases by mid-June (Figure 11). Although there could be missing data, the overall lower death rate could also be associated with the age composition of the affected people in Bangladesh. By mid-June, around 57% of the confirmed cases fall into the 21-40 age group, whereas age group 41-60+ constitutes 35% of the cases in Bangladesh. At the same time, Bangladesh’s percentage of the younger population aged between 10 and 40 is higher than that of the US, Italy, and France where most COVID-19 deaths constitute the 40+ age group. Nevertheless, Bangladesh is currently facing a deep humanitarian crisis as the broken health care and economic systems are not allowing treating all its citizens equally (Savage & Ahsan, 2020). Coordinated and targeted NPI measures could help significantly reduce transmission to the local community, as evidenced in the case of China where COVID-19 originated (Lai et al., 2020).

**Figure 10.**
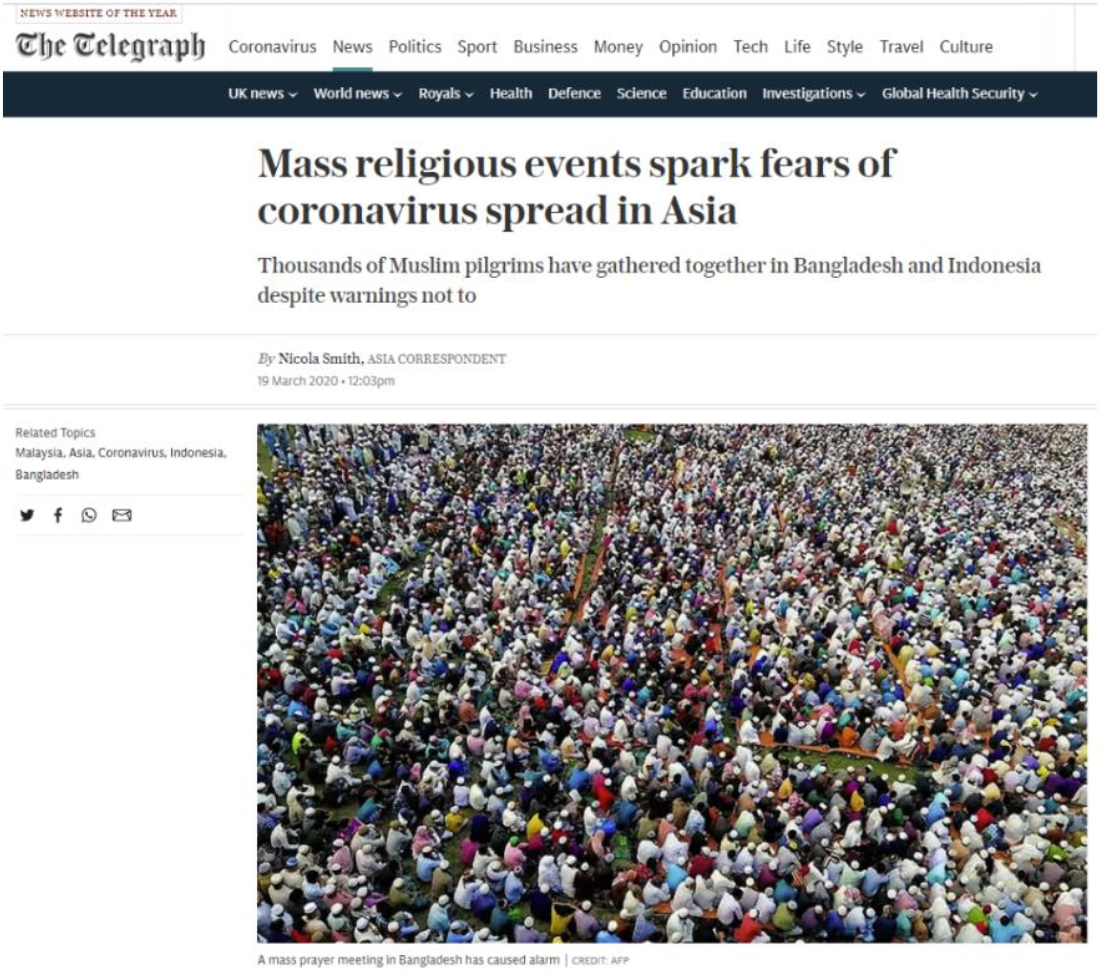
Mass congregation for ritual purpose at the early phase of the COVID-19 outbreak in Bangladesh (March 18)

**Figure 11.**
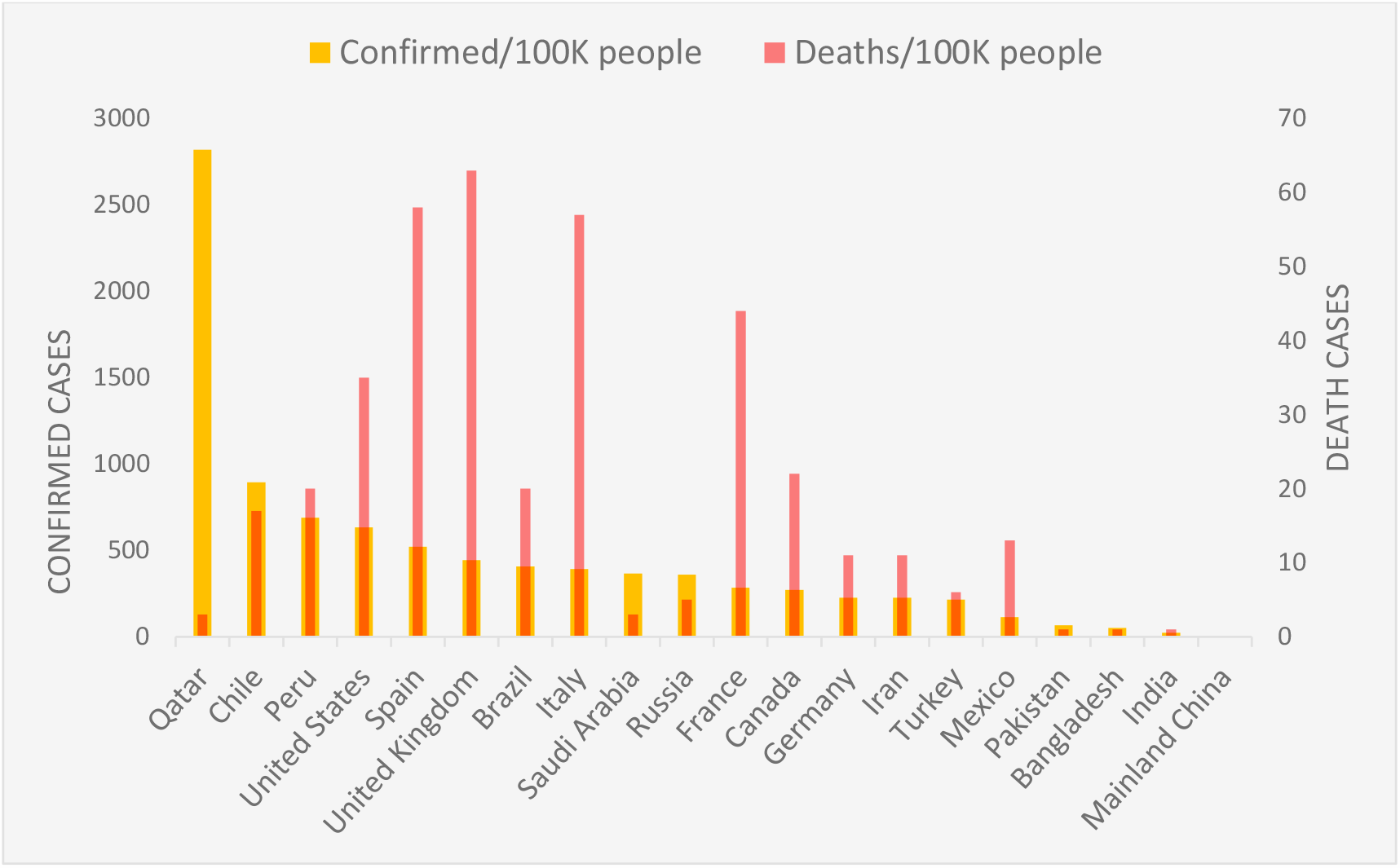
Number of COVID-19 confirmed and death cases per 100K people among the top 20 counties with the highest number of confirmed cases (up to June 14, 2020; Data source: CNN health by Pettersson, H., Manley, B., & Hernandez, S., 2020).

The timeliness and strength of this study lie in its contribution to rapid surveillance measures as manifested by the prospective space-time scan statistics approach. The prospective version of SaTScan is an excellent exploratory statistic approach for closely monitoring disease outbreaks (Kulldorff & Kleinman, 2015). However, some limitations can be addressed in future studies when more data become available. First, we only included confirmed cases reported by IEDCR. Inclusion of suspected and probable cases as additional covariates could improve the understanding of the true magnitude of risk. However, such data were not available or accessible at this point of time. Second, our analyses could not adjust for the age since age-specific data on COVID-19 cases was not available. However, experience worldwide has shown that COVID-19 can affect all age groups, although the recovery rate tends to vary among those groups (Desjardins et al., 2020). It could provide valuable insights into the space-time transmission rate and process through including age-specific data with other medical (pre-existing conditions) and socio-economic indicators (e.g. housing density, median income) at the local-level. Third, our findings may suffer from a low COVID-19 testing rate (513 per 1 million; WHO Bangladesh situation report, May 04, 2020) against its high density and size of the population in Bangladesh (Howlader & Khan, 2020; Nabi, M. S., 2020). However, as more missing and new data are added, the prospective scan statistic in this research will potentially detect new emerging clusters that would reveal different space-time transmission dynamics.

Overall, the space-time emerging patterns and change in the relative risks of the COVID-19 outbreak identified here can be further compared and validated with local-level epidemiological and travel network datasets. Thus, our study should be taken as a benchmark to understand and unraveling complex pattern and process of COVID-19 transmission in Bangladesh and elsewhere as more heterogeneous data become available and rigorous cross-disciplinary research is conducted in coming future. Meanwhile, knowledge derived in this study on Bangladesh highlights three specific guidelines for reducing disease transmission and minimizing impacts, particularly in the developing countries: i) in the face of a highly likely disease outbreak, act early to prevent or minimize infection cases by putting necessary travel restrictions, ii) coordinate with local civic and religious leaders to control human mobility – e.g. cultural or religious gatherings, sporting events, etc., and iii) take early preparations on the NPI measures as there are generally inadequate hospital and health care facilities in resource-scarce countries.

## Data Availability

Data sharing is not applicable to this article as no new data were created or analyzed in this study.

## Acknowledgements

We are grateful to Donna J. Peuquet for her constructive feedback that helped improving this manuscript. We also thank Monojit Saha for sharing some geospatial datasets used in this study.

## Funding

This research did not receive any funding grant from the public, commercial, or not-for-profit agencies.

## Appendix: A1

Distribution of Thana Population of 2018 in Dhaka megacity, Bangladesh

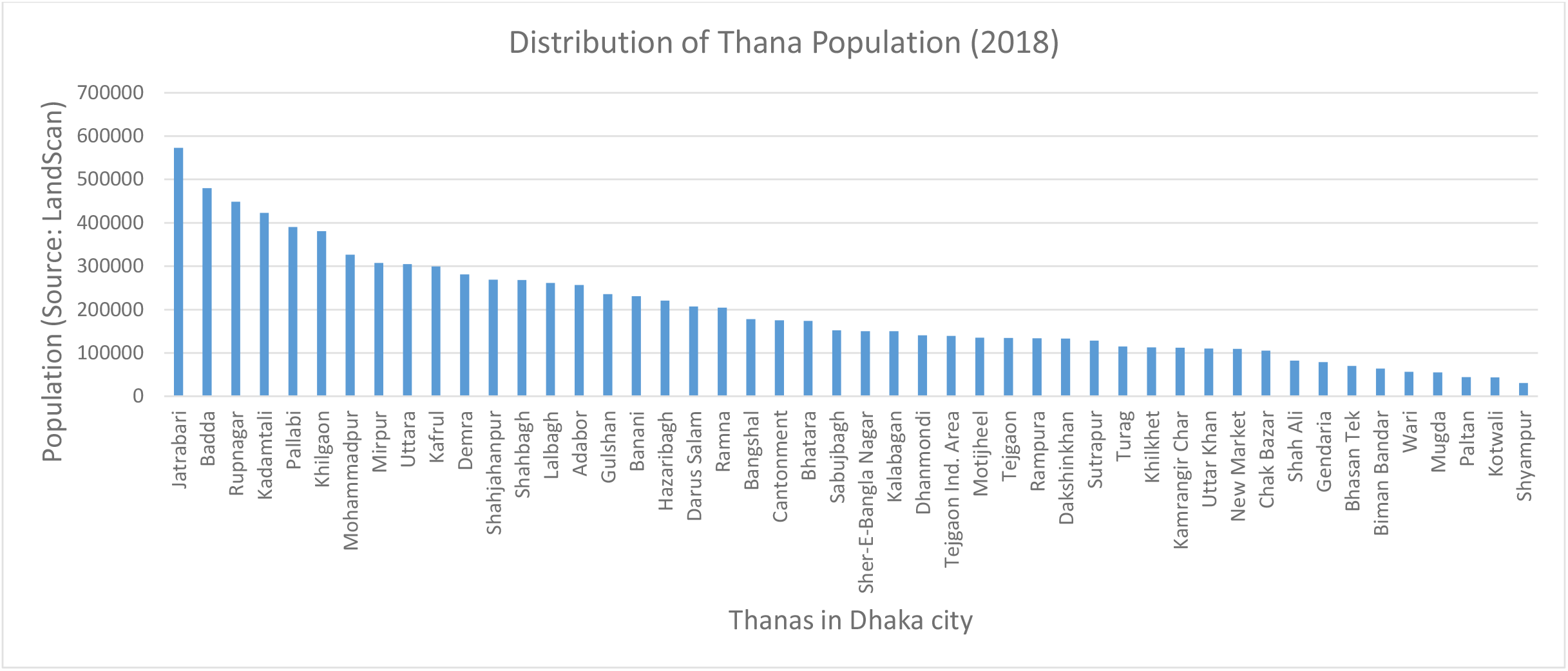

## Notes

### Competing Interest Statement

The authors have declared no competing interest.

### Author Declarations

This research does not include human subjects.

## References

Acter, T., Uddin, N., Das, J., Akhter, A., Rabia, T., & Kim, S. (2020). Evolution of severe acute respiratory syndrome coronavirus 2 (SARS-CoV-2) as coronavirus disease 2019 (COVID-19) pandemic?: A global health emergency. Science of the Total Environment, 730, 138996. https://doi.org/10.1016/j.scitotenv.2020.138996

Ahmed, A., & Rahman, M. M. (2020). COVID-19 trend in Bangladesh: deviation from epidemiological model and critical analysis of the possible factors, 1–14.

Bhuiyan, A. K. M. I., Sakib, N., Pakpour, A. H., Griffiths, M. D., & Mamun, M. A. (2020). COVID-19-Related Suicides in Bangladesh Due to Lockdown and Economic Factors: Case Study Evidence from Media Reports. International Journal of Mental Health and Addiction. https://doi.org/10.1007/s11469-020-00307-y

Chowdhury, R., Heng, K., Shawon, M. S. R., Goh, G., Okonofua, D., Ochoa-Rosales, C., … Global Dynamic Interventions Strategies for COVID-19 Collaborative Group. (2020). Dynamic interventions to control COVID-19 pandemic: a multivariate prediction modelling study comparing 16 worldwide countries. European Journal of Epidemiology, 35(5), 389–399. https://doi.org/10.1007/s10654-020-00649-w

Coleman, M., Coleman, M., Mabuza, A. M., Kok, G., Coetzee, M., & Durrheim, D. N. (2009). Using the SaTScan method to detect local malaria clusters for guiding malaria control programmes. Malaria Journal, 8(1). https://doi.org/10.1186/1475-2875-8-68

Delmelle, E., Casas, I., Rojas, J. H., & Varela, A. (2013). Spatio-temporal patterns of dengue fever in cali, Colombia. International Journal of Applied Geospatial Research, 4(4), 58–75. https://doi.org/10.4018/jagr.2013100104

Desjardins, M. R., Hohl, A., & Delmelle, E. M. (2020). Rapid surveillance of COVID-19 in the United States using a prospective space-time scan statistic: Detecting and evaluating emerging clusters. Applied Geography, 118(April), 102202. https://doi.org/10.1016/j.apgeog.2020.102202

Desjardins, M. R., Whiteman, A., Casas, I., & Delmelle, E. (2018). Space-time clusters and co-occurrence of chikungunya and dengue fever in Colombia from 2015 to 2016. Acta Tropica, 185(December 2017), 77–85. https://doi.org/10.1016/j.actatropica.2018.04.023

Dobson, J. E., Bright, E. A., Coleman, P. R., Durfee, R. C., & Worley, B. A. (2000). LandScan: A global population database for estimating populations at risk. Photogrammetric Engineering and Remote Sensing, 66(7), 849–857.

Dong, E., Du, H., & Gardner, L. (2020). An interactive web-based dashboard to track COVID-19 in real time. The Lancet Infectious Diseases, 20(5), 533–534. https://doi.org/10.1016/S1473-3099(20)30120-1

Ferreira, R. V., Martines, M. R., Toppa, R. H., Assunção, L. M., Desjardins, M. R., & Delmelle, E. M. (2020). Applying a Prospective Space-Time Scan Statistic to Examine the Evolution of COVID-19 Clusters in the State of São Paulo, Brazil. https://doi.org/10.1101/2020.06.04.20122770

Fong, M. W., Gao, H., Wong, J. Y., Xiao, J., Shiu, E. Y. C., Ryu, S., & Cowling, B. J. (2020). Nonpharmaceutical Measures for Pandemic Influenza in Nonhealthcare Settings-Social Distancing Measures. Emerging Infectious Diseases, 26(5), 976–984. https://doi.org/10.3201/eid2605.190995

Franch-pardo, I., Napoletano, B. M., Rosete-verges, F., & Billa, L. (2020). Spatial analysis and GIS in the study of COVID-19. A review. Science of the Total Environment, 739, 140033. https://doi.org/10.1016/j.scitotenv.2020.140033

Gatto, M., Bertuzzo, E., Mari, L., Miccoli, S., Carraro, L., & Casagrandi, R. (2020). Spread and dynamics of the COVID-19 epidemic in Italy?: Effects of emergency containment measures. https://doi.org/10.1073/pnas.2004978117

Giuliani, D., Dickson, M. M., Espa, G., & Santi, F. (2020). Modelling and predicting the spatio-temporal spread of Coronavirus disease 2019 (COVID-19) in Italy, (March).

Gross, B., Zheng, Z., Liu, S., Chen, X., Sela, A., Li, J., … Havlin, S. (2020). Spatio-temporal propagation of COVID-19 pandemics, 1–7.

Guan, W., Ni, Z., Hu, Y., Liang, W., Ou, C., He, J., … Zhu, S. (2020). Clinical Characteristics of Coronavirus Disease 2019 in China. The New England Journal of Medicine. https://doi.org/10.1056/NEJMoa2002032

Hohl, A., Delmelle, E., & Desjardins, M. (2020). Rapid detection of COVID-19 clusters in the United States using a prospective space-time scan statistic?: An update, 27–33.

Howlader, T., & Khan, H. R. (2020). Breaking the back of COVID-19 Is Bangladesh doing enough testing. Medrxiv, 1–13. https://doi.org/10.1101/2020.05.09.20096123

Howlader, T., Khan, H. R., & Islam, M. M. (2020). Battling the COVID-19 Pandemic Is Bangladesh Prepared. Medrxiv, 1–20. https://doi.org/10.1101/2020.04.29.20084236

Islam, M. T., Talukder, A. K., Siddiqui, M. N., & Islam, T. (2020). Tackling the Pandemic COVID-19?: the Bangladesh Perspective, (April), 1–18. https://doi.org/10.20944/preprints202004.0384.v1

Jones, R. C., Liberatore, M., Fernandez, J. R., & Gerber, S. I. (2006). Use of a prospective space-time scan statistic to prioritize shigellosis case investigations in an urban jurisdiction. Public Health Reports, 121(2), 133–139. https://doi.org/10.1080/08985620500531865

Kalam, A., & Hussain, A. M. (2020). Modeling and Analysis of The Early-Growth Dynamics of COVID-19 Transmission, (May), 1–26. https://doi.org/10.20944/preprints202005.0372.v1

Kamruzzaman, Md. “Bangladesh: Thousands Gather at Mosques amid Pandemic.” Anadolu Ajansı, 8 May 2020, www.aa.com.tr/en/asia-pacific/bangladesh-thousands-gather-at-mosques-amid-pandemic/1833947.

Kulldorff, M. (2018). SatScan user guide 2006, 8. Retrieved from www.satscan.org/

Kulldorff, Martin. (2001). Prospective time periodic geographical disease surveillance using a scan statistic. Journal of the Royal Statistical Society. Series A: Statistics in Society, 164(1), 61–72. https://doi.org/10.1111/1467-985X.00186

Kulldorff, Martin, Athas, W. F., Feuer, E. J., Miller, B. A., & Key, C. R. (1998). Evaluating cluster alarms: A space-time scan statistic and brain cancer in Los Alamos, New Mexico. American Journal of Public Health, 88(9), 1377–1380. https://doi.org/10.2105/AJPH.88.9.1377

Kulldorff, Martin, Heffernan, R., Hartman, J.,Assunção, R., & Mostashari, F. (2005). A space-time permutation scan statistic for disease outbreak detection. PLoS Medicine, 2(3), 0216–0224. https://doi.org/10.1371/journal.pmed.0020059

Kulldorff, Martin, & Kleinman, K. (2015). Comments on ‘A critical look at prospective surveillance using a scan statistic’ by T. Correa, M. Costa, and R. Assunção. Statistics in Medicine, 34(7), 1094–1095. https://doi.org/10.1002/sim.6430

Li, Q., Guan, X., Wu, P., Wang, X., Zhou, L., Tong, Y., … Feng, Z. (2020). Early transmission dynamics in Wuhan, China, of novel coronavirus-infected pneumonia. New England Journal of Medicine, 382(13), 1199–1207. https://doi.org/10.1056/NEJMoa2001316

Mollalo, A., Vahedi, B., & Rivera, K. M. (2020). GIS-based spatial modeling of COVID-19 incidence rate in the continental United States. Science of the Total Environment, 728, 138884. https://doi.org/10.1016/j.scitotenv.2020.138884

Monjur, M. R., & Hassan, M. Z. (2020). Early phases of COVID-19 management in a low-income country: Case of Bangladesh. Infection Control and Hospital Epidemiology, 2020. https://doi.org/10.1017/ice.2020.147

Orea, L., & Aacute;lvarez, I. C. (2020). How effective has the Spanish lockdown been to battle COVID-19? A spatial analysis of the coronavirus propagation across provinces.

Owusu, C., Desjardins, M. R., Baker, K. M., & Delmelle, E. (2019). Residential mobility impacts relative risk estimates of space-time clusters of chlamydia in Kalamazoo county, Michigan. Geospatial Health, 14(2), 254–264. https://doi.org/10.4081/gh.2019.812

Peeri, N. C., Shrestha, N., Rahman, M. S., Zaki, R., Tan, Z., Bibi, S., … Haque, U. (2020). The SARS, MERS and novel coronavirus (COVID-19) epidemics, the newest and biggest global health threats: what lessons have we learned? International Journal of Epidemiology, 1–10. https://doi.org/10.1093/ije/dyaa033

Rogerson, P. A. (1997). Surveillance systems for monitoring the development of spatial patterns. Statistics in Medicine, 16(18), 2081–2093. https://doi.org/10.1002/(SICI)1097-0258(19970930)16:18<2081::AID-SIM638>3.0.CO;2-W

Su, L., Hong, N., Zhou, X., He, J., Ma, Y., & Jiang, H. (2020). Evaluation of the secondary transmission pattern and epidemic prediction of COVID-19 in the four metropolitan areas of China, (Mcmc).

Tang, W., Liao, H., Marley, G., Wang, Z., Cheng, W., Wu, D., & Yu, R. (2020). The Changing Patterns of Coronavirus Disease 2019 (COVID-19) in China?: A Tempogeographic Analysis of the Severe Acute Respiratory Syndrome Coronavirus 2 Epidemic, 2019(Xx Xxxx), 1–7. https://doi.org/10.1093/cid/ciaa423

Wadood, M. A., Mamun, A., Rafi, M. A., Islam, M. kamrul, Mohd, S., Lee, L. L., & Hossain, M. G. (2020). Knowledge, attitude, practice and perception regarding COVID-19 among students in Bangladesh: Survey in Rajshahi University. MedRxiv Preprint. https://doi.org/10.1101/2020.04.21.20074757

Whiteman, A., Desjardins, M. R., Eskildsen, G. A., & Loaiza, J. R. (2019). Detecting space-time clusters of dengue fever in Panama after adjusting for vector surveillance data. PLoS Neglected Tropical Diseases, 13(9), 1–14. https://doi.org/10.1371/journal.pntd.0007266

Xiong, Y., Wang, Y., Chen, F., & Zhu, M. (2020). Spatial Statistics and Influencing Factors of the COVID-19 Epidemic at Both Prefecture and County Levels in Hubei Province, China. International Journal of Environmental Research and Public Health, 17(11).

